# A Sardinian founder mutation in *GP1BB* that impacts thrombocytopenia

**DOI:** 10.1101/2020.07.06.20143263

**Authors:** Fabio Busonero, Maristella Steri, Valeria Orrù, Gabriella Sole, Stefania Olla, Michele Marongiu, Carlo Sidore, Sandra Lai, Antonella Mulas, Andrea Maschio, Magdalena Zoledziewska, Matteo Floris, Mauro Pala, Paola Forabosco, Isadora Asunis, Maristella Pitzalis, Francesca Deidda, Marco Masala, Cristian Antonio Caria, Susanna Barella, Goncalo R. Abecasis, David Schlessinger, Serena Sanna, Edoardo Fiorillo, Francesco Cucca

**Affiliations:** Istituto di Ricerca Genetica e Biomedica, Consiglio Nazionale delle Ricerche (CNR), 09042 Monserrato (Cagliari), Italy; Dipartimento di Scienze Biomediche, Università degli Studi di Sassari, 07100 Sassari, Italy; Ospedale Pediatrico Microcitemico “Antonio Cao” (A.O.Brotzu), 09100 Cagliari, Italy; Center for Statistical Genetics, University of Michigan, Ann Arbor, 48109 Michigan, USA; Laboratory of Genetics and Genomics, National Institute on Aging, US National Institutes of Health, Baltimore, 21224 Maryland, USA

**Author notes:** These authors contributed equally to this work (Fabio Busonero, Maristella Steri). Correspondence should be addressed to Francesco Cucca, Fabio Busonero or Maristella Steri.

## Abstract

To investigate the genetic regulation of platelet (PLT) levels we carried out a whole-genome association analysis in 6,528 Sardinians from the general population of the Lanusei valley. We found 6 variants significantly influencing PLT levels, including a novel rare missense mutation (p.Pro27Ser) in the *GP1BB* protein that is associated with PLT reduction (P=1.17×10^−16^). This mutation is rare in the SardiNIA population cohort (frequency of 0.45%), even rarer in the rest of the Sardinian island (frequency of 0.16%), and not reported elsewhere. Notably, *GP1BB* is involved in Bernard-Soulier syndrome (BSS), a rare autosomal recessive bleeding disorder caused by a defect in the platelet GPIb-IX-V protein complex. Consistently, the 57 identified individuals heterozygous for the p.P27S mutation showed mild thrombocytopenia, morphologically enlarged platelets (P=2.13×10^−10^), and reduced expression of two GPIb-IX-V-complex components: GPIbα (−26.51%, P=3.66×10^−8^) and GPIX (−24.69%, P=2.66×10^−6^). Molecular modeling infers a corresponding reduction in the stability of GP1BB. These observations predict that in homozygosity as well as in individuals carrying specific compound heterozygous configurations, this variant likely causes BSS.

## Introduction

To date, Genome Wide Association Studies (GWAS) based on genotyped^1,2^ or imputed polymorphisms^3,4^ have identified numerous common, mostly regulatory, variants underlying signals associated with PLT levels. Moreover, whole-exome sequencing^5^ and whole-genome sequencing^6^ efforts (and derived Exomechip arrays^7,8^) allowed the investigation and detection of rare, mostly coding, variants associated with PLT levels. Overall, 301 independent loci have been identified as associated with PLT at the genome-wide significance threshold of 5×10^−8^, with effects on standard deviation ranging from 0.02 to 0.15 units and with minor allele frequencies from 0.09% to 50% (Suppl. Table 1)^9^. Reduction in PLT levels below a given threshold or qualitative platelets defects may result in diseases characterized by bleeding. The inherited platelets disorders are generally mild but can sometimes be severe, especially after trauma or surgical procedures, in some monogenic disorders. In particular, Bernard-Soulier syndrome (BSS; MIM #231200) is a rare (1 per 1 million live births) and often severe autosomal recessive thrombocytopenia and thrombocytopathy^10^. The hallmark of BSS is defective adhesion of platelets to the sub-endothelium, resulting from quantitative or qualitative defects in the GPIb-IX-V complex^11^, a platelet receptor for von Willebrand Factor (vWF), which is composed of four subunits: GPIbα, GPIbβ, GPIX, and GPV. Laboratory diagnosis is based on prolonged bleeding time, moderate-to-severe thrombocytopenia (platelet levels in affected individuals typically ranges from 20 to 100×10^9^/L), giant platelets on peripheral blood smears, and deficient ristocetin-dependent platelet agglutination (RIPA)^12^. However, very little is known about the biochemical and clinical features of heterozygous carriers of mutations causing BSS and about the impact in general population individuals of variation in genes encoding the GPIb-IX-V complex in heterozygosity. In fact, family members with only one mutated allele are generally asymptomatic, with sub-normal platelet number and function. Nevertheless, they might have slightly enlarged platelets and marginally reduced levels of GPIb-IX-V complex expression, as well as moderately reduced RIPA response. Here, a sequencing-based whole-genome association analysis was performed in 6,528 general population individuals who are part of the SardiNIA study^13^, identifying a number of variants significantly influencing platelet levels. They include a novel founder non-synonymous mutation (22:19711445:C/T) in the *GP1BB* gene, defects in which are involved in BSS.

## Subjects and Methods

### Cohorts and sample description

The SardiNIA project^13^ is a longitudinal cohort study of over 8,000 general population volunteers, recruited in the Lanusei Valley, phenotyped for thousands of quantitative traits, including PLT, MPV and other hematological traits, often underlying clinical endpoints for common and rare diseases. All participants gave informed consent to study protocols, which were approved by the Sardinian local research ethic committees: Ethical Committee of ASSL of Lanusei (2009/0016600) and Ethical Committee of ASSL of Sassari (2171/CE).

### Blood platelet levels and volume measurement

PLT levels were measured in 6,528 volunteers, while mean platelet volume (MPV) was assessed in a subset of 2,000 of them. Blood samples were collected for each volunteer, and divided into two aliquots; the first used for genomic DNA extraction and the second to characterize several blood phenotypes, including PLT and MPV using the Beckman COULTER LH750 Series Hematology Systems, according to manufacturer’s instructions. Over the time, several instruments have been changed, so that PLT and MPV were also measured with Beckman Coulter LH700 2727, and LH750 3435 series. To avoid biases due to different instruments, these were considered as a covariate (1 for the LH700 and 2 for LH750). Given the International System (SI) unit, these physiological ranges for number of circulating platelets and volume, were considered: 150-450×10^9^/L, and 8-13 femtoliter, respectively.

### Genotyping and imputation

Genetic analyses were performed using a genetic map based on 6,602 samples genotyped with four Illumina arrays (OmniExpress, ImmunoChip, Cardio-MetaboChip and ExomeChip) as previously described^14^. Imputation was performed on a genome-wide scale using the Sardinian sequence-based reference panel of 3,514 individuals and the *minimac* software on pre-phased genotypes. After imputation, only markers with imputation quality (RSQR)>0.3 for MAF>=1% or RSQR>0.6 for MAF<1% were retained for association analyses^13^, yielding ∼22 million variants (20,143,392 SNPs and 1,688,858 indels) useful for analyses.

### Association analysis

Before performing GWAS, PLT and MPV phenotypes were both inverse-normalized and adjusted by sex, age, age^2^, smoking status, aspirins assumption and instrument employed for measurements, as covariate. In the same model, the effect on traits of the body mass index was also tested but it wasn’t included in the analyses being not significant. Additive effects were searched using EPACTS (epacts-3.2.6 version)^15^, a software that implements a linear mixed model adjusted with a genomic-based kinship matrix calculated using all quality-checked genotyped SNPs with MAF>1%. The advantage of this model is that the kinship matrix encodes a wide range of sample structures, including both cryptic relatedness and population stratification. As a proof of appropriate adjustment of all confounders, the genomic inflation factor (lambda GC) for PLT GWAS was 0.988. To identify independent signals, a conditional GWAS analysis for each trait was performed by adding the leading SNPs found in the primary GWAS as covariates to the basic model. A SNP reaching the standard genome-wide significance threshold for sequencing-based GWAS (P<6.9×10^−9^) was considered as significant^13^. To calculate the heritability explained by each variant the following formula was used:

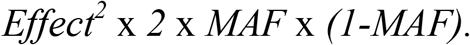

### Gene-prioritization strategy on 22q11.21 associated locus

Statistical fine-mapping of the 22q11.21 associated region was performed by calculating the 95% credible set, i.e. the minimum set of variants having a 95% summed posterior probability of containing the causal variant. The credible set was calculated using FINEMAP v1.3 and setting at most one causal variant (−-n-causal-snps 1). To predict the functional effects and pathogenicity of the 10 candidate variants in the credible set, several *in silico* methods were considered: Polymorphism Phenotyping v2 (PolyPhen-2), Sorting Intolerant From Tolerant (SIFT), Mutation Taster, and CADD-score annotations included in VEP^16^.

### Platelet staining protocol and Immunophenotyping

Fresh blood samples were collected in Vacutainer tubes containing sodium citrate as anticoagulant. Cell phenotyping was carried-out by flow-cytometry immediately after blood collection to avoid any time-dependent artifacts. Blood samples were then diluted 1:20 with phosphate-buffered saline (PBS, 20 μL of blood and 800 μL of PBS), stained with the antibody mix (Suppl. Table 2) for 20 minutes in refrigerated condition, fixed with 1% formalin, then analyzed by FACS Canto II and Diva software (BD Biosciences). All antibodies were obtained from BD Biosciences. Moreover, to dissect platelet activation status, samples were incubated with adenosine diphosphate (ADP, final concentration 20 μM)^17^ for 5 minutes at room temperature, then stained as described for basal conditions.

### Flow-cytometry data analyses

Statistical analyses of the cytometric experiments were performed using the R software (R version 3.4.3 (2017-11-30)^18^. The Shapiro-Wilk Normality test was used to verify whether the fluorescence intensities were normally distributed. Markers expression was normalized by CD61 values, to take into account differences in platelet size between p.P27S carriers and controls, while statistical significance was assessed with the two-sided Mann-Whitney test (also known as unpaired two-samples Wilcoxon test). For both CD42a (GPIX) and CD63 (Granulophysin) markers, one volunteer for each of the two experimental groups (C/C and C/T) were discarded from the analyses given their outlier antigen expression values.

### Molecular modeling analysis

Molecular dynamics study was performed on the wild type (WT) and mutated (p.P27S) protein using the 3D structure of glycoprotein Ib-β available on Protein Data Bank (PDB code 3RFE)^19^. The structure of p.P27S protein has been carried out using pymol software. NAMD software package^20^ was used to perform all-atom molecular dynamics MD and using CHARMM force field^21,22^. The VMD program was used for the preparation and analysis phase of simulations. In details, the two structures were solvated in TIP3P water box of water with 12 Å buffering distance. Assuming normal charge states of ionizable groups corresponding to pH 7, sodium Na^+^ andchloride Cl^−^ ions at physiological concentration of 0.15 mol/L were added to achieve charge neutrality, more closely mimicking a realistic biological environment. Systems were subjected to minimization for 10,000 steps, equilibration for 1 ns and dynamics simulations lasting 100 ns.

During minimization and equilibration all C α atoms of the complex were restrained with a one Kcal/mol/Å^2^. Root-Mean-Square-Fluctuations (*RMSF*) were calculated for WT and p.P27S proteins. Two software tools, Strum^23^ and SDM^24^, were used to quantify WT and p.P27S proteins stability, while STRIDE^25^ and SDM were employed to calculate the solvent accessibility. Electrostatic surface potentials were calculated using APBS method^26^. Individual charges were assigned using pdb2pqr software^27^ with the CHARMM force field. Final images were generated with VMD from -10kT to 10kT.

## Results

Interrogating 21,832,250 variants, either directly genotyped or successfully imputed, 5 reported genetic associations with PLT at the *ARHGEF3*^1^, *HBS1L*^1^, *GCSAML*^3^, *TP53BP1*^4^ and *TPM4*^1^ loci were replicated in population-based GWAS^13^ (Suppl. Table 3, and Supplementary material for description). In addition, a novel association in the 22q11.21 region was found (Suppl. Fig. 1). The novel signal encompassed numerous rare variants in linkage disequilibrium (LD) spanning 1 Mb (Fig. 1A). The variant with the lowest P-value (22:19940476:A/T, effect=-1.186 s.d., P=3.428×10^−17^) fell in an intron of the *COMT* gene. Association at this variant was completely independent of previously reported associations in the same genomic region (see Suppl. material). Considering the predicted functional impact of 10 variants in the 95% credible set of candidates (Suppl. Table 4), only one coding variant (22:19711445:C/T, MAF=0.0045, r^2^=0.924 with top variant), mapping in the second exon of the *GP1BB* gene (c.C79T, p.P27S), was considered for downstream functional evaluation. No homozygous and 57 carriers for 22:19711445:C/T rare allele were found and further validated by Sanger sequencing. Platelet levels in wild-type homozygous were 242.87±117.05×10^9^/L (mean±1.96*SD), whereas in p.P27S carriers were 174.17±91.51×10^9^/L, corresponding to a reduction of 70.13×10^9^ platelet/L for each copy of the minor allele (Fig. 1B). With this large effect, the novel mutation explains about 1.05% of phenotypic variance (h2) for PLT levels, representing the largest phenotypic effect among all the independent variants reported so far in the GWAS Catalog (Suppl. Table 5)^9^. Moreover, in a subset of 2,000 individuals, whose mean platelet volume (MPV) was measured, this variant was significantly associated (P=2.13×10^−10^; Fig. 2A) with notably larger platelets (10.62±1.20 fL in carriers; 9.14±0.98 fL in wild-type homozygotes). This finding is consistent with clinical evidence of morphologically enlarged defective platelets in thrombocytopenic patients, as in BSS (Fig. 2B). To verify whether the p.P27S variant in *GP1BB* gene shows different frequencies in the cohort of the Lanusei valley compared to the rest of the island, we further genotyped it in 1,235 whole-genome sequenced individuals from across Sardinia^13^. Four additional heterozygous individuals were found, resulting in a lower MAF (0.00162); according to Hardy-Weinberg expectation, about 4 homozygous BSS cases are predicted in the island. By contrast, p.P27S is completely missing in large sequencing datasets such as UK10K^7^, 1,000 Genomes Project (1,000GP)^8^, GoNL^28^, GnomeAD browser Broad Institute (126,216 exomes and 15,137 genomes)^29^, and the Exome Sequencing Project (ESP) data^30^ in NHLBI’s TOPMed program^31^.

**Figure 1.**
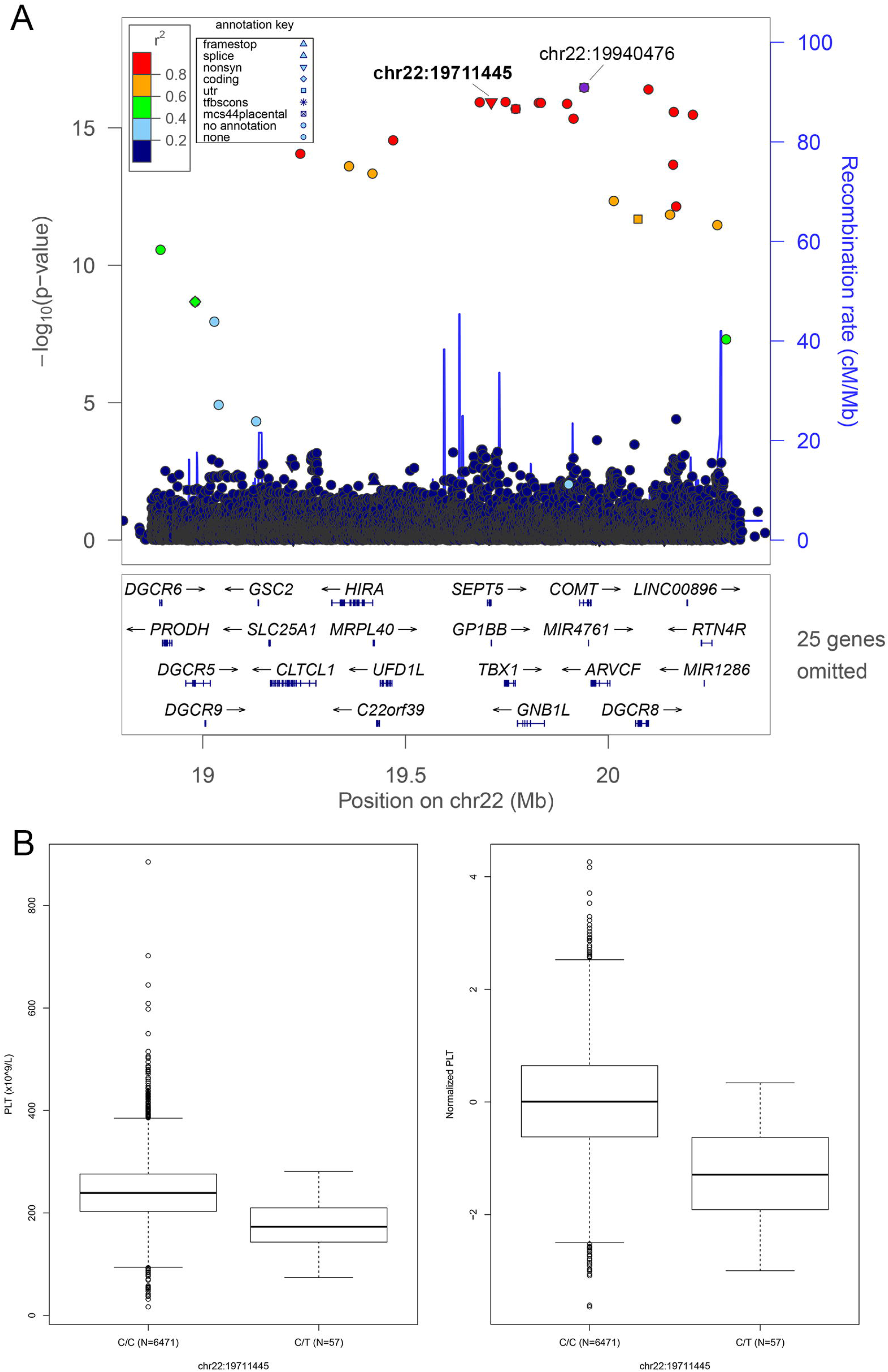
Regional association plot and box plot for the top signal in GP1BB. Shown in ***A***, are association results in the *GP1BB* region for platelet level using the Sardinian sequencing-based reference panel. The association strength (expressed as - log_10_ P values; *y* axis) is plotted versus the genomic positions (on the hg19/GRCh37 genomic build; *x* axis) around the most significant SNP chr22:19940476, indicated with a purple dot. Other variants in the region are color-coded to reflect their extent of linkage disequilibrium with the top variant (taken from pairwise r2 values calculated on Sardinians haplotypes) The missense variant chr22:19711445 is also indicated. Symbols reflect genomic functional annotation, as indicated in the related inner box. The right y-axis corresponds to recombination rate (cM/Mb), plotted as a solid blue line overlying the plot. The box at the bottom shows specification on genes and exons, as well as the direction of transcription, according to RefSeq^39^. This plot is made using the standalone version of LocusZoom package^40^. Shown in ***B***, are box plots of platelet levels (***left***) and normalized platelet levels (***right***) distribution, stratified according to chr22:19711445 genotype for 57 heterozygous carriers and 6,471 homozygous wild types.

**Figure 2.**
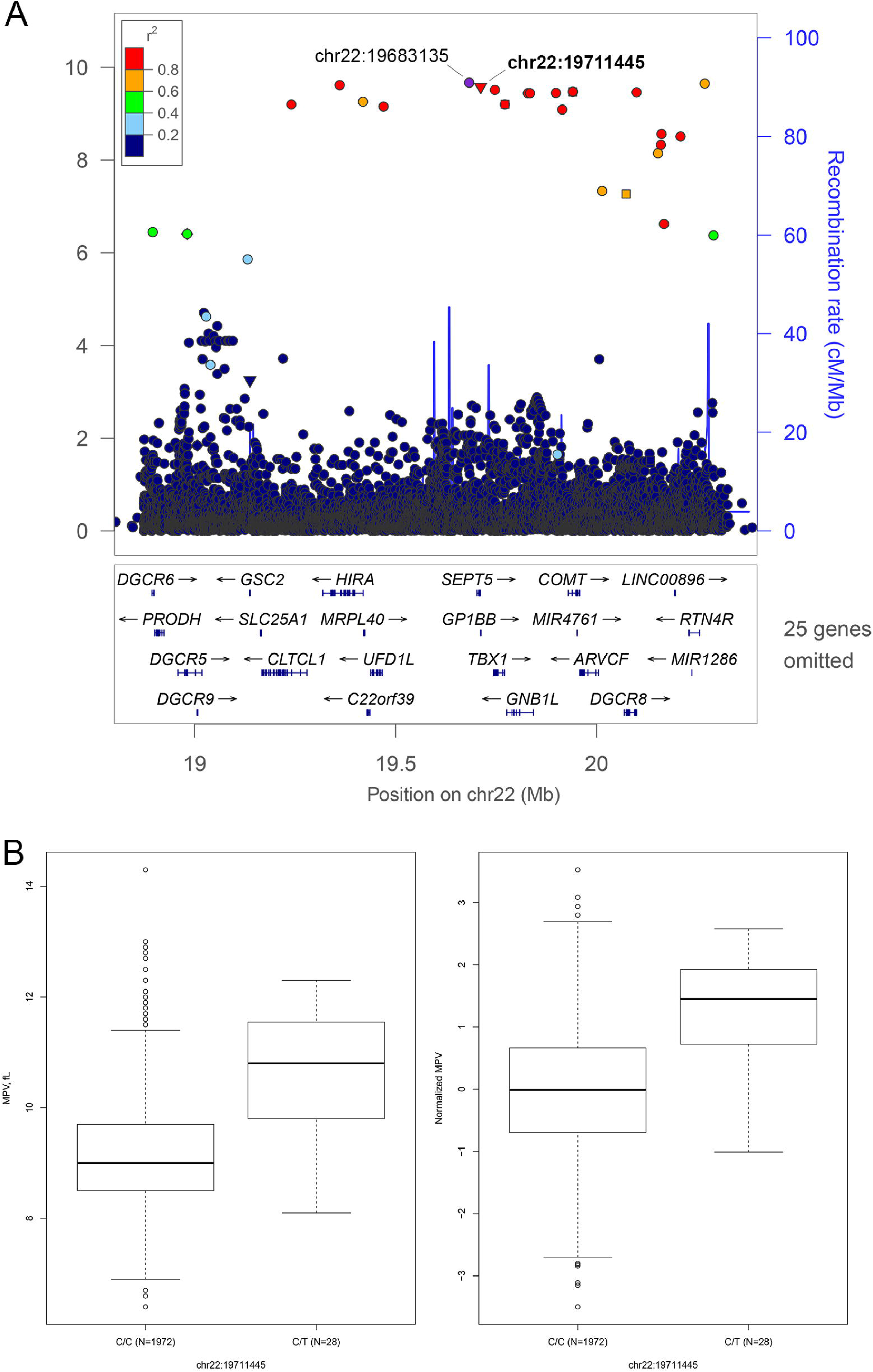
Regional association plot and box plots for the top signal in GP1BB. Shown in ***A***, are association results in the *GP1BB* region for mean platelet volume using the Sardinian sequencing-based reference panel. The association strength (expressed as - log_10_ P values; *y* axis) is plotted versus the genomic positions (on the hg19/GRCh37 genomic build; *x* axis) around the most significant SNP chr22:19683135, indicated with a purple dot. See ***Fig. 1*** legend for detailed description. Shown in ***B***, are the box plots of mean platelet volume **(*left*)** and normalized platelet volume **(*right*)** distribution, stratified according to chr22:19711445 genotype for 28 heterozygous carriers of and 1,972 homozygous wild types.

### Functional study of p.P27S mutation

Functional consequences and the likely mechanisms of action of p.P27S mutation were investigated by flow cytometry and molecular modeling analysis.

### A. p.P27S mutation affects basal expression level of GPIX and GPIbα glycoproteins

Platelet functionality was assessed by a 7-color flow cytometry panel (Suppl. Table 2) in 24 of 57 p.P27S carriers (42.1%) and in an equal number of matched unrelated controls. Monoclonal antibodies (MoAbs) directed against the GPIIb-IIIa (integrin αIIbβ3) complex (CD41a/PCy7 and CD61/FITC), and the von Willebrand factor receptor complex (CD42a/eFluor450, CD42b/PE) were used to investigate the basal receptor expression in resting platelets. Indeed, since the GPIIb-IIIa complex is constitutively expressed on platelets and megakaryocytes, CD41a and CD61 are both used in clinical diagnosis of BSS (Suppl. Fig. 2A). Platelets of p.P27S carriers showed increased levels of integrins GPIIb (CD41a, +22.36%, Mann-Whitney test P=1.61×10^−4^, N=48) and GPIIIa (CD61, +16.20%, P=6.61×10^−4^, N=48) in the αIIbβ3 complex - a typical finding in the presence of enlarged platelets (Suppl. Fig. 2B and 2C). The expression of GPIX and GPIbα glycoproteins and their correct assembly into the GPIb-IX-V complex are known to be impaired by a defective GPIbβ peptide^32,33^. Indeed, compared to controls, despite carrying only one mutated allele, in basal conditions p.P27S heterozygous showed appreciably lower expression levels of both GPIX (−24.69%, P=2.66×10^−6^, N=46; Fig. 3A) and GPIbα (−26.51%, P=3.66×10^−8^, N=48; Fig. 3B), and consequently less of the entire complex. This is far more than reported in a recent report^34^, which described normal expression levels of GPIX and GPIbα in carriers of other missense mutations in *GP1BB*.

**Figure 3.**
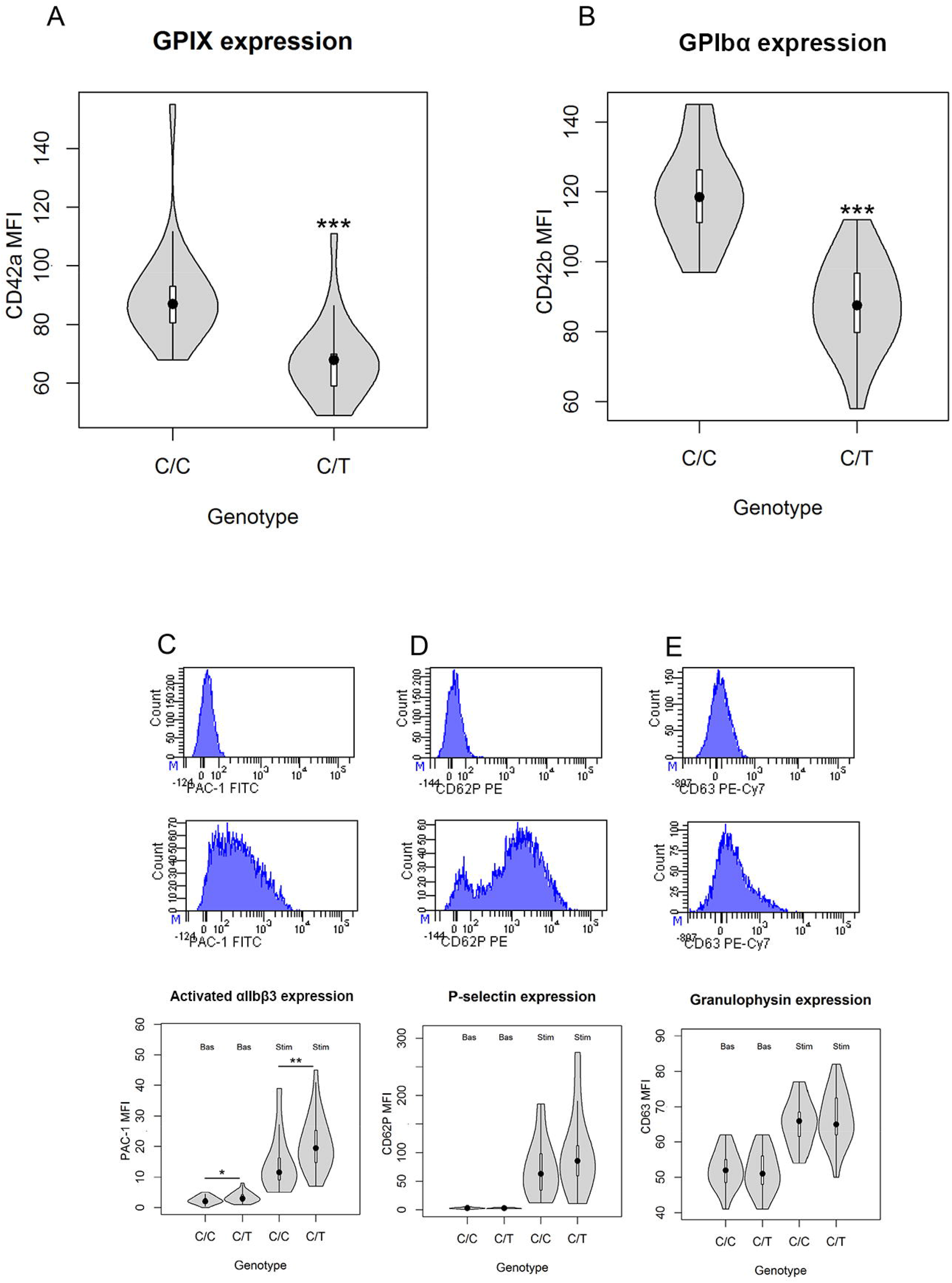
Evaluation of platelet function by flow-cytometry. Shown are basal expression levels of the main GPIb-IX-V receptor glycoproteins, GPIX (***A***) and GPIbα (***B***) on resting platelets. For each genotype class, chr22:19711445 heterozygous carriers are compared to homozygous wild types. For GPIX 23 carriers and 23 controls were assessed, while for GPIbα 24 carriers and 24 controls were available. Moreover, a representative sample of change in the most relevant platelet activation-dependent markers (PAC-1, CD62P and CD63), in basal condition and after stimulation with ADP, along with their expression level changes, is shown (***C, D, E***, respectively). *p<0.05, **p<0.01, ***p<0.001.

### B. Platelet activation is differentially modulated in p.P27S carriers

Platelet activation results in a complex series of changes, including physical redistribution of receptors, variation in their molecular conformation, and granule content secretion^17^ reflecting platelet function. To investigate pre-activation and reactivity changes between p.P27S in carrier and control platelets, the expression of several activation-markers (high-affinity conformation of αIIbβ3, P-selectin, granulophysin) on the platelet surface was assessed after exposure to the agonist adenosine diphosphate (ADP). Activated integrin αIIbβ3 was prominently induced in p.P27S carriers compared to controls, as shown by the extent of PAC-1 binding to resting and activated platelets (+41.94%, P=4.84×10^−3^, N=48, Fig. 3C). This pronounced effect of the allele contrasts with the unexpected finding of an unaltered response of platelets recently observed after ADP and collagen stimulation in both BSS patients and carriers^32^. Remarkably, platelet reactivity turned out to be differentially regulated: in fact, after ADP induction, no changes were observed in surface exposure of neo P-selectin (CD62P, +35.22%, P=0.138, N=48) and neo granulophysin (CD63, +1.86%, P=0.658, N=46, Fig. 3D-E), markers of granule content release.

### C. GPIb-IX-V complex assembly and stability is affected by p.P27S mutation

The unique functional effects of the p.P27S lead us to examine its possible consequences on molecular structure and conformational changes of GPIbβ. Indeed, the effect of the amino-acid change in glycoprotein Ib-β was tested by molecular modeling analysis based on the x-ray crystal structure^19^ and building a model for p.P27S substitution.

*GP1BB* encodes a 206 amino acid long transmembrane protein. The Proline to Serine substitution fell in the Leucine-rich repeat N-terminal (LRRNT) domain (Figg. 4A-B for 2-D and 3-D structures, respectively). Proline residues are expected to be disruptive of structure; and indeed, in that highly conserved region and close to cysteine residues involved in the Cys26-Cys32 disulphide bridge in the structure, p.P27S could thus modify the stability, and consequently the conformation, of GPIbβ. To test this hypothesis, we first performed *in-silico* Molecular Dynamic (MD) simulations. An increased conformational mobility of the amino acid backbone was indeed indicated close to p.P27S (Suppl. Fig. 3), suggesting instability of the GPIbβ glycoprotein. This is in accord with the observed reduction of the level of GPIbα and GPIX glycoproteins. Moreover, a greater fluctuation was also observed in p.P27S compared to WT protein, as indicated by Root-Mean-Square-Fluctuation (RMSF; Suppl. Fig. 3); this, in turn, is reflected in a major fluctuation of the amino acids in loop 2 of the p.P27S protein (Fig. 4C).

**Figure 4.**
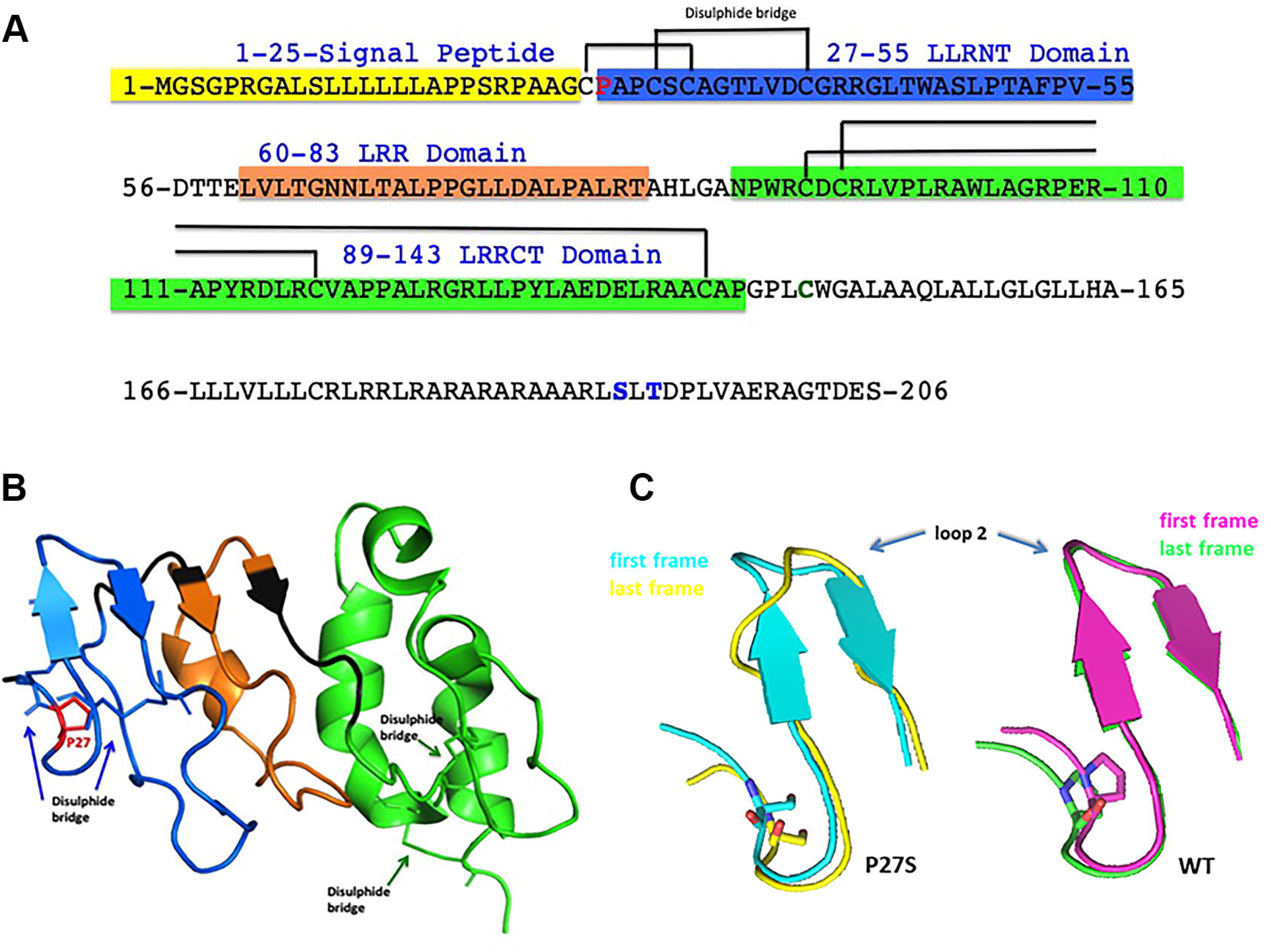
Molecular modeling analyses. Shown is the 2-D representation of platelet glycoprotein (GP)Ibβ sequence (***A***), with highlighted the signal peptide (yellow), the LRRNT domain (blue), the LRR domain (orange) and the LRRCT domain (green). Shown in red is the Proline27 to Serine27 substitution, the C147 (green) forming a disulphide bridge with GP1bα, and the phosphorylated amino acids S191, T193 (blue). Shown is the 3-D structure of platelet glycoprotein (GP)Ibβ sequence (***B***), colour-coded according to the legend of the 2-D representation. Shown is the X-ray structure of protein (26-143 aa code 3RFE). Shown is the impact of p.P27S on GPIbβ glycoprotein conformation (***C***). The superposition between the first (pink) and last (green) frame of the molecular dynamics for the WT protein (***left***), and the superposition between the first (***teal***) and last (***yellow***) frame of the molecular dynamics for the p.P27S protein are represented.

The reduction of protein stability due to p.P27S was corroborated with two software tools to calculate differences in free energy between the WT and mutated protein (ddG=-1,35 and ddG=-0.66, respectively^23,24^)(Fig. 5A). Furthermore, p.P27S mutation caused a slight increase in solvent accessibility, as calculated with two software tools: 13% for Pro and 18,6% for Ser, 16.3 Å^2^ for Pro and 20.7 for Ser, respectively^24,25^) as well as changes in the electrostatic potential surface (Fig 5B) that likely affects the conformation suitable for normal platelet function, especially influencing the optimal protein-protein interaction for the correct GPIb-IX-V complex assembly.

**Figure 5.**
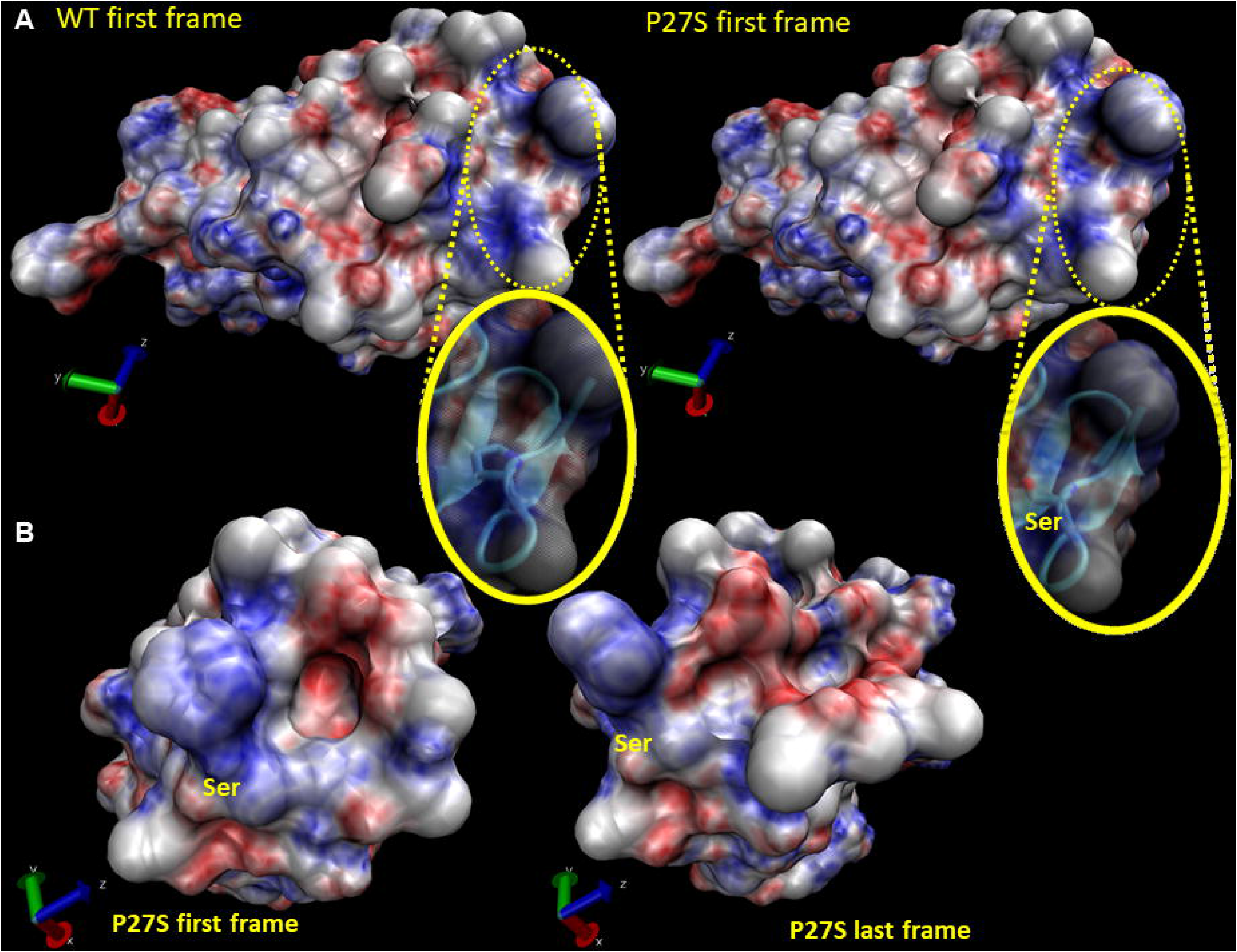
Electrostatic surface potential calculation. Shown are the electrostatic surface potentials ***(A)*** of the first frame of the WT (***left***) and p.P27S protein (***right***). Negative and positive charges are indicated in red and blue, respectively (−10kT and 10kT). In ***B***, the first (***left***) and last frames (***right)*** of the mutated protein (***B***).

## Discussion

Sequencing-based genome-wide association efforts in founder and genetically isolated populations, through the identification of associations with variants that are very rare or absent elsewhere, help enlarge the spectrum of biochemical, molecular and clinical signatures due to genetic variants. Here, to dissect the impact of genetic variability on platelet-related traits in Sardinia, a whole-genome association study in 6,528 volunteers of the SardiNIA general population cohort was carried-out. Six signals were identified, including a novel Sardinian-specific missense mutation (22:19711445:C/T; NM_000407, exon 2, c.C79T, p.P27S) in *GP1BB* gene. All typical findings of macrothrombocytopenias (i.e. BSS) were observed in p.P27S obligate carriers characterized in this study: low levels of large platelets and low expression of GPIX and GPIbα glycoproteins, as shown by flow-cytometry experiments.

As one might anticipate, the most severe cases are caused by deletions and nonsense mutations, but some missense mutations are disabling enough to be clinically significant. In one of the reported cases^35^, a charge difference is introduced (p.Asn89Asp); in the other^36^, as in this case, the Proline residue is replaced (p.Pro27Leu), which is expected to disrupt secondary structure, in the protein sequence. That the Proline to Serine substitution at position 27 influences conformational changes and stability of the GPIbβ glycoprotein, in turn affecting GPIb-IX-V complex function, is further clearly supported by the *in-silico* molecular dynamic analyses. vWF receptor complex 1:2:1 stoichiometry consisting of 4 GPIbβ glycoproteins covalently-bonded with 2 GPIbα glycoproteins and non-covalently paired with 2 subunits each of GPIX and GPV, may explain why a single mutated allele in GPIbβ significantly affects the function of this complex^11^. In fact, a critical interaction of GPIbβ with GPIX glycoprotein involves N-terminal residues 15 through 32 of GPIbβ, precisely including Proline 27^37^. And indeed, only when coexpressed with GPIbβ can GPIX be detected on the cell surface, again indicating that GPIbβ interacts with and stabilizes GPIX.

From these findings, at least 4 homozygous individuals for the p.P27S mutation, most likely with BSS, are expected in Sardinia, but none has been reported: this may suggest that BSS is likely underdiagnosed in Sardinia, consistent with reports from other studies^38^. Clinicians should thus be aware of the novel p.P27S mutation in the molecular characterization of patients born in Sardinia or of Sardinian origin with a clinical picture of platelet macrocytosis and PLT levels <20-100×10^9^/L, and therefore at high-risk of cerebral hemorrhage.

## Data Availability

https://sardinia.irp.nia.nih.gov/Background/background.html

## Authorship

F.B., M.Pi. and F.D. collected samples and extracted genomic DNA from blood; F.B., A.M. and M.Z. performed genotyping; F.B., A.Ma., performed sequencing; V.O. and E.F. designed flow cytometric panels; V.O., E.F. and S.L. performed cytometric analysis; M.S., G.S., C.S., M.F., M.Pa., P.F., Ma.Ma. and S.S. performed statistical analyses; M.S., M.M. and S.S. performed bioinformatic analyses; F.B. and S.S. performed region-specific analysis and selected candidate genes; S.O. performed in-silico analyses; I.A. and C.A.C. performed other functional evaluation; F.C., G.R.A and D.S. provided funds and supervised the work; S.B. provided clinical support; F.B., M.S. and F.C. wrote the paper; V.O., G.S., S.O., C.S., A.Ma., M.F., D.S., and S.S. revised the paper. All authors read the paper and contributed to its final form.

## Acknowledgements

We thank all the volunteers who generously participated in this study; we are grateful to mr Mario Lovicu and mr Nazario Olla for the logistic support provided and helpful suggestions. Supported by contracts N01-AG-1-2109 and HHSN271201100005C from the Intramural Research Program of the National Institute on Aging, National Institutes of Health (NIH).

## Web Resources

The URLs for data presented herein are as follows

- UK10K, https://www.uk10k.org/;
- 1000 Genomes Project data repository, ftp://ftp.1000genomes.ebi.ac.uk/;
- GWAS catalog, https://www.ebi.ac.uk/gwas/;
- SardiNIA Project, https://sardinia.irp.nia.nih.gov/;
- Online Mendelian Inheritance in Man, https://www.omim.org/
- GoNL, http://www.nlgenome.nl/;
- GnomeAD, http://gnomad.broadinstitute.org/;
- Exome Sequencing Project, https://esp.gs.washington.edu/drupal/
- HRC, http://www.haplotype-reference-consortium.org/;
- Protein Data Bank, https://www.rcsb.org/;
- Minimac, http://genome.sph.umich.edu/wiki/Minimac/;
- EPACTS, http://genome.sph.umich.edu/wiki/EPACTS/;
- Variant Effect Predictor, https://www.ensembl.org/Tools/VEP;
- Pymol, www.Pymol.org/;
- RefSeq, https://www.ncbi.nlm.nih.gov/refseq/;
- LocusZoom package, http://locuszoom.org/;

